# Distinct exosomal miRNA profiles from BALF and lung tissue from COPD and IPF patients

**DOI:** 10.1101/2021.08.24.21262557

**Authors:** Gagandeep Kaur, Krishna P Maremanda, Michael Campos, Hitendra S Chand, Feng Li, Nikhil Hirani, M.A. Haseeb, Irfan Rahman

## Abstract

**Background:** Chronic Obstructive Pulmonary Disease (COPD) and Idiopathic Pulmonary Fibrosis (IPF) are chronic, progressive lung ailments which are characterized by distinct pathologies. Early detection biomarkers and disease mechanisms for these debilitating diseases are lacking. Exosomes are small extracellular vesicles attributed to carry proteins, mRNA, miRNA and sncRNA to facilitate cell-to-cell communication under normal and diseased conditions. Exosomal miRNAs have been studied in relation to many diseases. However, there is little to no knowledge regarding the miRNA population of BALF or the lung tissue derived exosomes in COPD and IPF. Here, we determined and compared the miRNA profiles of BALF and lung tissue-derived exosomes from healthy non-smokers, healthy smokers, and patients with COPD and IPF in independent cohorts.

**Results:** Exosome characterization using NanoSight particle tracking and TEM demonstrated that the BALF-derived exosomes were approximately 89.85 nm in size and ∼2.95 × 10^10^ particles/mL. Lung-derived exosomes were ∼146.04 nm in size and ∼2.38 × 10^11^ particles/mL. NGS results identified three differentially expressed miRNAs in the BALF, while one in the lung-derived exosomes from COPD patients as compared to healthy non-smokers. Of these, three- and five-fold downregulation of miR-122-5p amongst the lung tissue-derived exosomes from COPD patients as compared to healthy non-smokers and smokers, respectively. Interestingly, there were key 55 differentially expressed miRNAs in the lung tissue-derived exosomes of IPF patients compared to non-smoking controls.

**Conclusions:** Overall, we identified specific miRNAs to develop as biomarkers or targets for pathogenesis of these chronic lung diseases.

## Introduction

Tobacco smoking remains the most prevalent preventable cause of morbidity and mortality, worldwide. Comprising of more than 5000 compounds (1), cigarette smoke is the leading risk-factor for developing chronic obstructive pulmonary disease (COPD) and idiopathic pulmonary fibrosis (IPF) in humans. Despite their distinct clinical features, both COPD and IPF can be defined as chronic, progressive airway diseases associated with increased risk of cancer development (2, 3). The current therapies for these conditions are mainly palliative; and the chief reason of this is due to limited understanding of the pathophysiology of these respective ailments (4, 5).

Evidence from literature suggest the role of extracellular vesicles/exosomes in the disease severity and outcome in COPD and IPF (6-10). Exosomes function to maintain homeostasis and intracellular stability. However, they also become pathosomes due to harmful stimulus (e.g. tobacco smoke) and can participate in the progression of diseases. In this respect, exosomes are known to cause pathological changes including oxidative stress, chronic inflammation, apoptosis, aging, epigenetic alterations and multi-organ dysfunction in COPD (11-14). Interestingly, exosomes are produced and released in the sputum, serum and BALF of COPD patients in large quantities which makes them a useful target to develop non-invasive diagnostics in COPD. Previous studies have mostly compared the serum-derived exosome populations from COPD patients and healthy individuals (15-20). Similarly, exosomes isolated from the biological fluids cause pro-inflammatory responses in lung cells (11, 21, 22). However, there is little to no knowledge about the BALF or the lung tissue-derived exosome populations in COPD or IPF.

Based on this, we compared the miRNA population in the BALF and lung tissue-derived exosomes from healthy non-smokers, healthy smokers, and patients with COPD and IPF in several independent cohorts. Numerous studies have shown that circulating miRNAs are involved in the progression, development and severity of various diseases including COPD and IPF (6, 9, 11, 23-26). These are also considered to be known targets for biomarker development (27, 28). Hence, we compared the exosome-derived miRNA profiles amongst COPD and IPF patients with healthy individuals to identify miRNA signatures that might be unique to each of these distinct pathological conditions and help determine the progress of the pulmonary damage at an early stage.

## Materials and Methods

### Ethics/Approval

The human patients and the patients’ data included in the study were procured from several agencies (described below) as human subjects were not directly involved in this work. The procurement of human lung tissues and BALF samples as deidentified samples was approved by the Materials Transfer Agreement and Procurement (Institutional Review Board, IRB), and Laboratory protocols by the Institutional Biosafety Committee (IBC) at the University of Rochester Medical Center, Rochester, NY. The project codes and dates of approval were as follow: Project Code: DRAI1 001 Protocol: 004, Date of approval and IRB/IBC approvals 2/11/2017 and 9/29/2017.

All the procedures/ protocols were carried per the guidelines and regulations specified by the University of Rochester, Rochester, NY. Other approvals include: (a) IRB study number 20080326 at the University of Miami, and (b) registered clinical trial (NCT04016181) and ethically approved by the University of Edinburgh (07/S1102/20) and NHS Lothian 2007/R/RES/02 by 14/06/2007. Additional samples were obtained from baseline measurements of Feasibility of Retinoids for the Treatment of Emphysema (FORTE) trial participants as described previously (29, 30).

### Study population and Sample Collection

We employed bronchoalveolar lavage fluid (BALF) and lung tissues collected from healthy (Non-smokers and Smokers) and diseased (COPD and IPF) human subjects as samples for this study from 7 independent cohorts (**Table 1**). A total of 40 BALF samples and 32 lung tissue samples were chosen for this study from multiple sources. The majority of the BALF samples used in this study were procured from a commercial provider-BioIVT (Westbury, NY, USA). Rest of the BALF samples were provided by our collaborators-Dr. Michael Campos from Division of Pulmonary, Allergy, Critical Care at University of Miami, Dr. Hitendra Chand from Department of Immunology at Florida International University, Dr. Haseeb Siddiqi from Department of Cell Biology at SUNY Downstate Health Sciences University and Dr. Nikhil Hirani from Center of Inflammation research at Edinburgh University, UK. The samples procured from our collaborators were validated for their disease categories based on their spirometry and clinical status.

**Table1:**
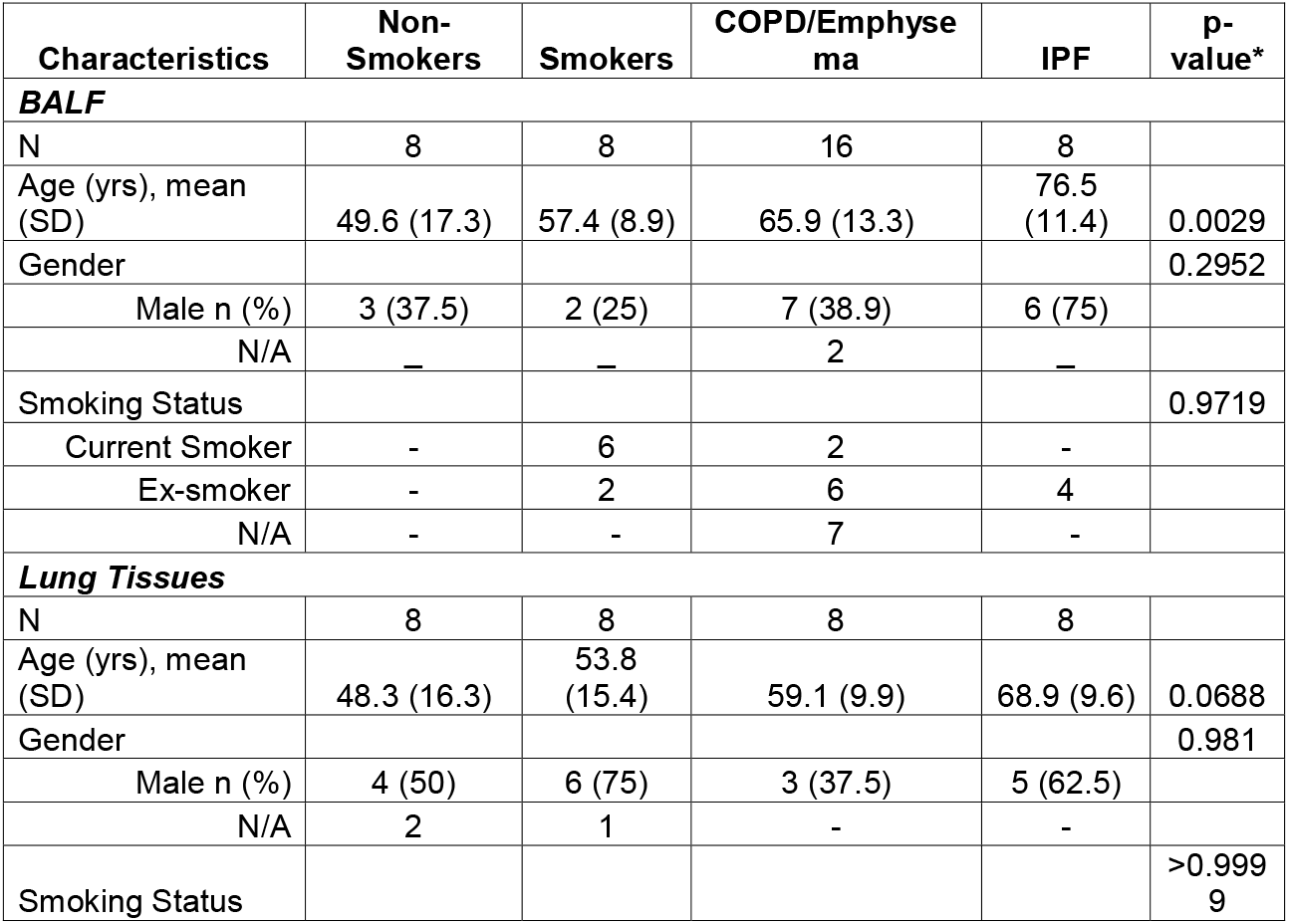
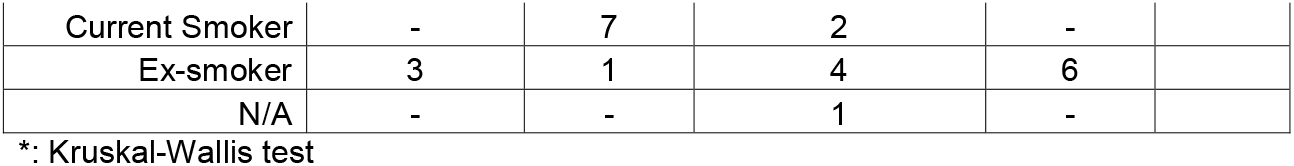
Clinical characteristics of study subjects.

| Characteristics | Non-Smokers | Smokers | COPD/Emphysema | IPF | p-value* |
| --- | --- | --- | --- | --- | --- |
| **BALF** |  |  |  |  |  |
| N | 8 | 8 | 16 | 8 |  |
| Age (yrs), mean (SD) | 49.6 (17.3) | 57.4 (8.9) | 65.9 (13.3) | 76.5 (11.4) | 0.0029 |
| Gender |  |  |  |  | 0.2952 |
| Male n (%) | 3 (37.5) | 2 (25) | 7 (38.9) | 6 (75) |  |
| N/A | — | — | 2 | — |  |
| Smoking Status |  |  |  |  | 0.9719 |
| Current Smoker | - | 6 | 2 | - |  |
| Ex-smoker | - | 2 | 6 | 4 |  |
| N/A | - | - | 7 | - |  |
| **Lung Tissues** |  |  |  |  |  |
| N | 8 | 8 | 8 | 8 |  |
| Age (yrs), mean (SD) | 48.3 (16.3) | 53.8 (15.4) | 59.1 (9.9) | 68.9 (9.6) | 0.0688 |
| Gender |  |  |  |  | 0.981 |
| Male n (%) | 4 (50) | 6 (75) | 3 (37.5) | 5 (62.5) |  |
| N/A | 2 | 1 | - | - |  |
| Smoking Status |  |  |  |  | >0.9999 |
| Current Smoker | - | 7 | 2 | - |  |
| Ex-smoker | 3 | 1 | 4 | 6 |  |
| N/A | - | - | 1 | - |  |

\*: Kruskal-Wallis test

Likewise, the lung samples were procured from three sources; (a) commercially available resource for procurement of human tissue and organ-NDRI (National Disease Research Interchange), (b) NHLBI-funded bio-specimen repository-LTRC (Lung Tissue Research Consortium), and (c) Department of Medicine and Pathology at the University of Helsinki Hospital, Finland as reported previously (31, 32).

All the subjects included in the study were above 21 years of age. Care was taken to include equal numbers of males and females in each subject group. A detailed characteristic of the BALF and Lung tissue samples used for this study is provided in **Table 1**.

### BALF exosome isolation

We employed commercially available Plasma/Serum Exosome Purification and RNA Isolation Midi Kit from Norgen Biotek (Cat# 58500; Ontario, Canada) to isolate exosomes from human BALF samples. BALF exosomes were isolated as per manufacturer’s protocol. In brief, 1 mL BALF sample was mixed with Nuclease-free water, ExoC Buffer and Slurry E; and incubated for 5 minutes at room temperature. Next, the solution was centrifuged at 2000 rpm for 5 min at room temperature and the supernatant was discarded. The slurry pellet was then resuspended in ExoR buffer and incubated for 10 min at room temperature. Thereafter the suspension was centrifuged at 8000 rpm for 2 min duration at room temperature and transferred to Mini Filter Spin column to elute the exosomal fraction. The eluted exosomes were then stored at −80°C until further use.

### Lung Tissue Exosome Isolation

The tissue exosomes were isolated using the protocol described by Dooner et al (2018 (33) with some modifications. In brief, 30-40 mg of lung tissue was chopped and lysed using 1X Liberase solution containing 0.01% DNase. The tube containing tissue lysate was left on an orbital shaker at 37°C for 1-hour duration to allow complete digestion of lung tissues. After 1-hour incubation, the tissue lysate was collected. The eluate was then centrifuged at 300 g for 10 min at 4°C to remove cell debris. Next, the supernatant was transferred to fresh tube and centrifuged at 2000 g for 10 min at 4°C. Again, the supernatant was transferred to a fresh tube and centrifuged at 10,000 g for 30 min at 4°C to remove larger vesicles. After this, the supernatant was transferred to ultracentrifuge tubes and the exosomes were pelleted at 110,000g for 70 min at 4°C using Optima Max-XP ultracentrifuge (Beckman Coulter, Brea, CA). At this stage, the supernatant was discarded and the pellet was resuspended in 1X PBS prior to filtering through 0.22µM filter. The filtrate was once again spun at 110,000g for 70 min at 4°C. Finally, the supernatant was discarded and the pellet was re-suspended in 1mL 1X PBS. This contained freshly isolated tissue exosomes that were stored at −80°C for future analysis.

### Exosome Characterization

We employed Hitachi 7650 Analytical transmission electron microscopy (TEM) to visualize the isolated exosomes and nanoparticle tracking analysis (NanoSight NS300) to analyze the particle size and concentration as described earlier (12, 34).

We also used immunoblotting to identify exosomal markers from the isolated fraction to characterize the BALF and lung tissue derived exosomes. In brief, 20 ug of exosomal lysate was resolved on a 10% sodium dodecyl sulfate (SDS)-polyacrylamide gel and electroblotted onto nitrocellulose membranes. Membrane was blocked using 5% blocking buffer for 1 hr and thereafter probed overnight with antibodies for exosomal surface markers. The antibodies include-CD9 (Cat# ab92726), CD63 (Cat# ab134045) (Abcam, Cambridge, UK) and CD81 (Cat# EXOAB-CD81A-1) (SBI Biosciences, Palo Alto, CA). The following days, the blots were washed and probed with appropriate secondary antibodies. Chemiluminescence was detected on the Bio-Rad ChemiDoc MP, Imaging system using the SuperSignal West Femto Maximum Sensitivity Substrate (Cat# 34096, Thermo Scientific, Waltham, MA).

### Exosomal RNA extraction

Total RNA from BALF exosomes was isolated using Exosomal RNA isolation kit (Norgen Bioteck Corporation, Cat# 58000) as per the manufacturer’s protocol. The detailed procedure has been published earlier (34).

Alternately, we used miRNeasy Mini Kit (Cat# 217004, Qiagen, Hilden, Germany) to isolate RNA from lung exosomes as per manufacturer’s protocol. Briefly, 700 µl of QIAzol lysis buffer was mixed with 250 µl of exosomal fraction and the mix was homogenized using QIAshredder. The homogenate was then mixed with 140µl of chloroform to allow phase separation and the aqueous phase was transferred to a fresh tube. Thereafter, the RNA was precipitated using 100% ethanol and washed using RWT and RPE buffers provided with the kit. Finally, the RNA was eluted using RNase-free water and stored at −80°C until further use. The RNA quality and quantity were checked using Agilent 2100 Bioanalyzer.

### Library preparation

The isolated RNA samples were shipped to Norgen Biotek, Canada for library preparation, sequencing and data analyses. The library preparation was performed using the standard library preparation workflow of Norgen including 3’ and 5’ adapter ligation, followed by reverse transcription, indexing PCR and size selection using a 6% Novex TBE Gel. In brief, Norgen Biotek Small RNA Library Prep Kit (Cat# 63600) was employed for library preparation making sure to use the same lot between each batch of samples.

Samples were quantified using both PicoGreen and Bioanalyzer. 6uL of high-quality total RNA was mixed with 3’ adaptor and T4 RNA ligase 2 to set up a reaction for 3’ Adaptor ligation per manufacturer’s protocol. This was followed by the removal of excess 3’ Adaptor and then 10-12 µl of final eluate was mixed with 5’Adaptor to set up a reaction for 5’ Adaptor Ligation. Next, the reaction for cDNA synthesis was set using the obtained ligated product as input, per manufacturer’s directions and incubated at 50°C for 1 hour in a thermocycler. This was followed by PCR amplification and indexing as advised and cleanup of final indexed PCR product using NGS Reaction Cleanup Kit. After cleaning, the samples were run on a 6% Novex TBE Gel for 50 minutes at 140V. The adaptor dimer not containing any library was excised, and the sample was eluted from the gel and checked for quality as per the manual’s instruction. At this stage, the library quality check was performed to estimate library size and concentration using Bioanalyzer. Samples were then pooled in equimolar ratios and further size selected using a 3% Agarose Gel cassette on the Pippin prep (Part # SAG-CDP3010). The pool was quantified by Bioanalyzer before starting the Next Generation Sequencing (NGS) run.

### Next Generation Sequencing (NGS) and Data Analysis

We employed NextSeq 500/550 High Output Kit v2 for 75 cycles (Cat# 20024906, Illumina, San Diego, CA) to perform NGS on our pooled library. Per the manufacturer’s directions the pooled library was denatured and diluted to the required concentration of 20pM for optimal cluster generation. Library was then applied onto the suitable flow-cell and sequenced using Illumina NextSeq 500 sequencing platform.

The raw sequence reads were analyzed by the team of bioinformaticians at Norgen using their advanced analysis pipeline for the processing of raw counts and alignment to endogenous genome and annotated transcriptome.

### Gene Ontology and KEGG analyses

The gene ontology or GO (35) enrichment analysis was performed, through the examination of significant GO terms associated with the differentially expressed miRNAs for each comparison group. The analysis was performed by iteratively testing the enrichment of each annotated GO term correlated with the set of pre-selected differentially expressed genes (in our case, miRNAs) in a linear fashion. Individual enriched annotated GO terms were analyzed using a Fisher’s exact test for both up-regulated and down-regulated genes in which GO terms with an FDR adjusted p-value threshold of 0.05 were reported as significantly relevant. The FDR is the false discovery rate generated using the Benjamini-Hochberg method, which adjusts the p-value based on the FDR. The analysis was performed separately on all three GO domains, i.e, biological process, molecular function and cellular component.

The KEGG (36) enrichment analysis was also performed to identify the differentially expressed genes within an associated pathway for various biological processes. The analysis was performed by testing the enrichment of each biological pathway with the associated gene (or miRNA) found within the set of pre-selected differentially expressed genes. Individually enriched pathways were then contrasted and compared between the two test groups using a Fisher’s exact test for both up-regulated and down-regulated genes within the pre-selected set of differentially expressed genes. Biological pathways with an adjusted p-value below 0.05 were reported.

### Statistical analysis

The miRNA data from various batches were normalized using trimmed mean of M-values (TMM) normalization method (37). The TMM normalized read counts were used for differential expression analysis. The Principal Component analysis (PCA) was plotted using the *ggfortify* function in R-software (version: 3.5.1) to produce a sample clustering plot based on miRNAs with the highest variation across all samples. The coefficient of variation (% CV) was calculated based on the log_2_ of TMM normalized data and then the 50 miRNAs with the highest %CV were selected and used to generate the PCA plot. The highest two components of variation were plotted on the x-axis (the first principal component, PCA1) and the y-axis (the second principal component, PCA2). Confidence ellipses and average center points were calculated and added for each sample group to further organize the biological groupings.

EdgeR statistical software package was used for DE analysis as described previously (38, 39). The Benjamini-Hochberg procedure was then used for adjusting the false discovery rate (FDR) (40). This allowed identifying the significant DE when comparing two groups. The DE was considered significant if log fold change of ≥ 1 or ≤ −1 at p-value and FDR of ≤ 0.05 was reported for the miRNA target. We used the *ggplot2* function in R software (version: 3.5.1) to plot volcano plots for illustrating a large number of miRNAs and displaying the particular miRNA targets with statistically significant differential expression.

Heat maps were generated using the *ComplexHeatmap* function in R-software (version: 3.5.1). The coefficient of variation (% CV) was calculated based on log_2_ of TMM normalized data and then the 50 miRNAs with the highest %CV were selected and used to generate the heat map.

Kruskal-Wallis test was used to calculate significance for sample distribution.

## Results

### Characterization of BALF and lung-tissue derived exosomes

Exosomes are known to be involved with intercellular communication thus affecting the physiological processes in various tissues. Here, we analyzed the miRNA population from the BALF and lung tissue exosomes isolated from the non-smokers, smokers, and the patients with COPD or IPF. We first isolated the BALF and lung tissue-derived exosomes using the methods described earlier. We employed immunoblotting, nanoparticle tracking analysis (NTA: NanoSight 300), and transmission electron microscopy (TEM) to characterize the isolated exosomes. We first used NTA to determine the particle concentration, size, or distribution of exosomes in isolated samples from BALF and lung tissues. The lung-derived exosomes (avg. conc. = 2.38 □ 2.2 × 10^11^ particles/mL) had larger size (mean = 146.04 nm). On the other hand, the average size of the BALF-derived exosomes was ∼89.85 nm (avg. conc. = 2.95 □ 2.2 × 10^10^ particles/mL) (Figs 1i & 2i). TEM analysis confirmed the morphology of isolated exosomes from BALF and lung tissue samples as shown in Figs 1ii & 2ii.

**Figure 1:**
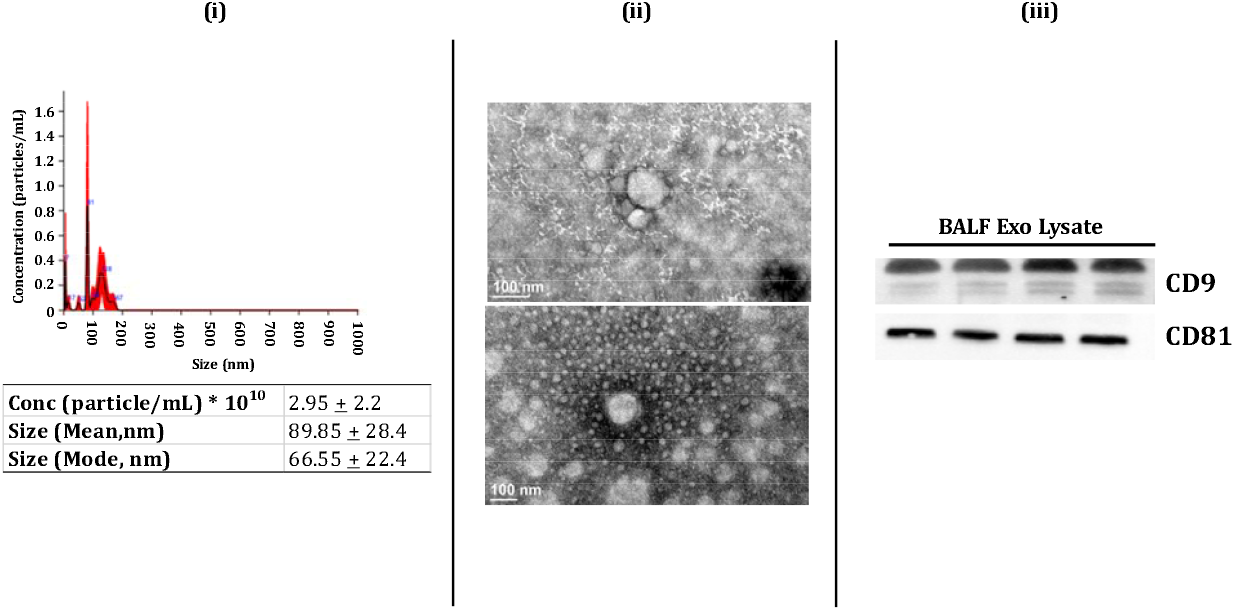
Characterization of human BALF-derived EVs/Exosomes. (i) Particle size depicted as mean, mode, and particle concentration were estimated using NanoSight NS300 (n=3-8/group). (ii) Representative TEM images of BALF-derived EVs/Exosomes (n=3). (iii) Immunoblot analysis of positive (CD9 and CD81) exosomal markers derived from human BALF (n=4).

**Figure 2:**
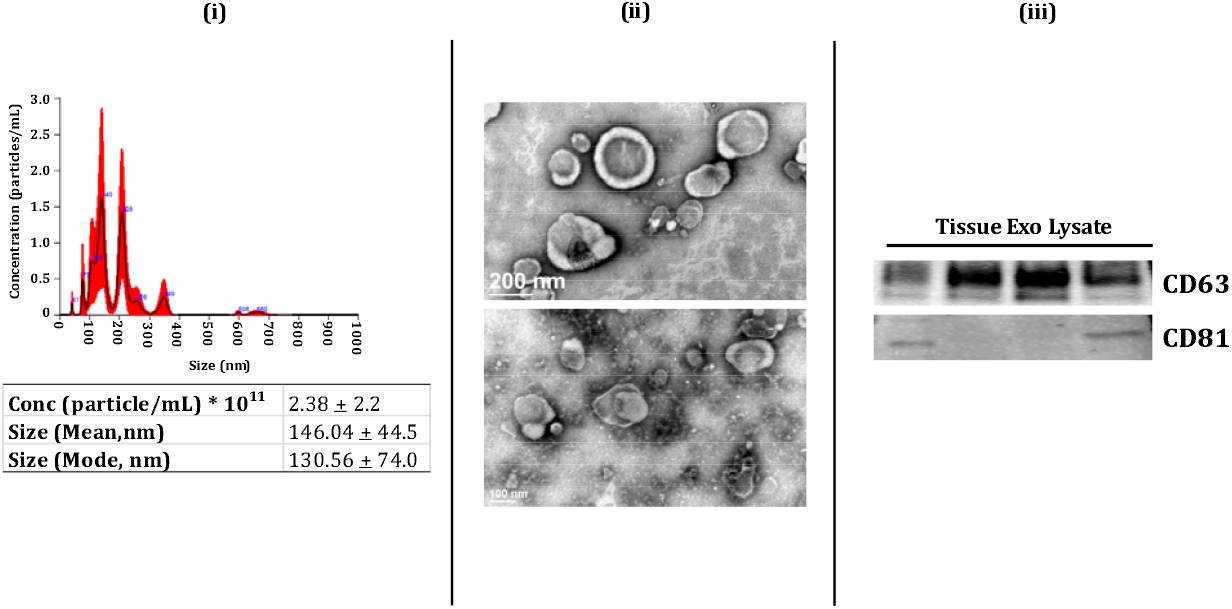
Characterization of human lung tissue-derived EVs/Exosomes. (i) Particle size depicted as mean, mode, and particle concentration were estimated using NanoSight NS300 (n=3-5/group). (ii) Representative TEM images of lung tissue-derived EVs/Exosomes (n=6). (iii) Immunoblot analysis of positive (CD63 and CD81) exosomal markers derived from human lung tissue (n=4).

Finally, we used immunoblotting to study the presence of exosome surface markers (CD9, CD81 and CD63) in the isolated exosome fractions from BALF and lung tissues. We found enrichment of positive surface markers for BALF exosomes, such as CD9 and CD81 in the isolated exosome fractions (Fig 1iii, full blots in Suppl. Fig 1). Similarly, we found abundance of positive surface markers for tissue exosomes-CD63 and CD81-in the exosome fractions from the lung tissues (Fig 2iii, full blots in Suppl. Fig 2). Overall, our results confirm the successful isolation of exosomes from human BALF and lung tissues in our study groups.

### Batch variations in the exosome-derived miRNA expression profiles amongst the various study groups

We performed Principal Component analyses (PCA) to visualize the batch variations within the samples. Separate analyses were run for the BALF- and lung tissue-derived exosomes. The plot was generated by using 50 miRNAs with the highest component of variation among groups. Each sample group was clustered using a confidence ellipse as shown in the Fig 3. The PCA plot from lung-derived exosomal miRNAs showed a distinct clustering of the IPF patient samples as compared to the other three study groups, thus suggesting a unique transcriptomic identity of these lung-derived exosomes.

**Figure 3:**
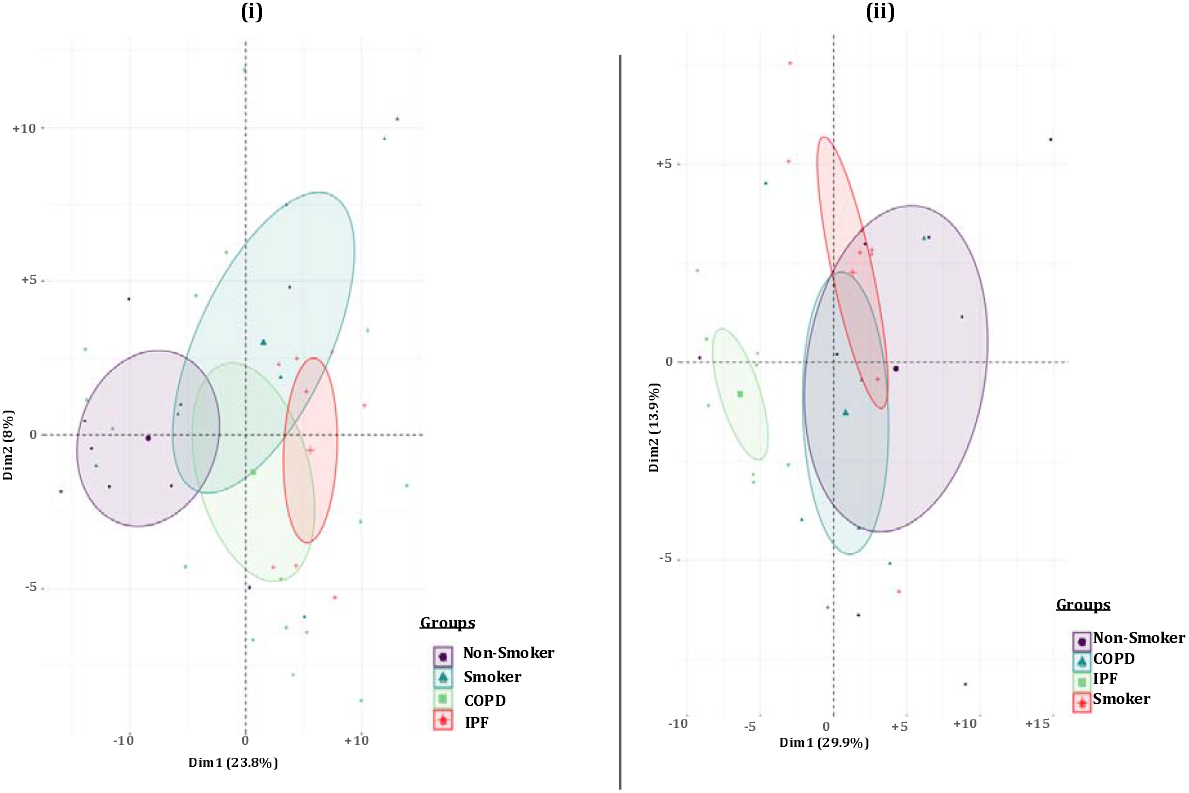
Principal Component Plot. Principal Component Analyses based on differential microRNA expression in individual (i) BALF- and (ii) Lung tissue-derived exosome samples from non-smokers, cigarette smokers, COPD and IPF subjects.

### Pairwise comparison of BALF- and lung tissue-derived exosomal miRNAs expression profiles

Next, we generated volcano plots showing pairwise comparisons of the differential miRNA expression profiles between various experimental groups in BALF or lung tissue-derived exosomes (Figs 4&5). We plotted the -log_10_ of adjusted *p*-values on the Y-axis, and the log_2_ fold change between two experimental groups on the X-axis to generate a volcano plot. Fold changes greater than ±1 on the logarithmic (base2) scale of thus derived volcano plots were considered significant. miRNAs showing no significant fold change were denoted with blue, while significantly up- or down-regulated miRNAs were denoted with green and red colored dots respectively.

**Figure 4:**
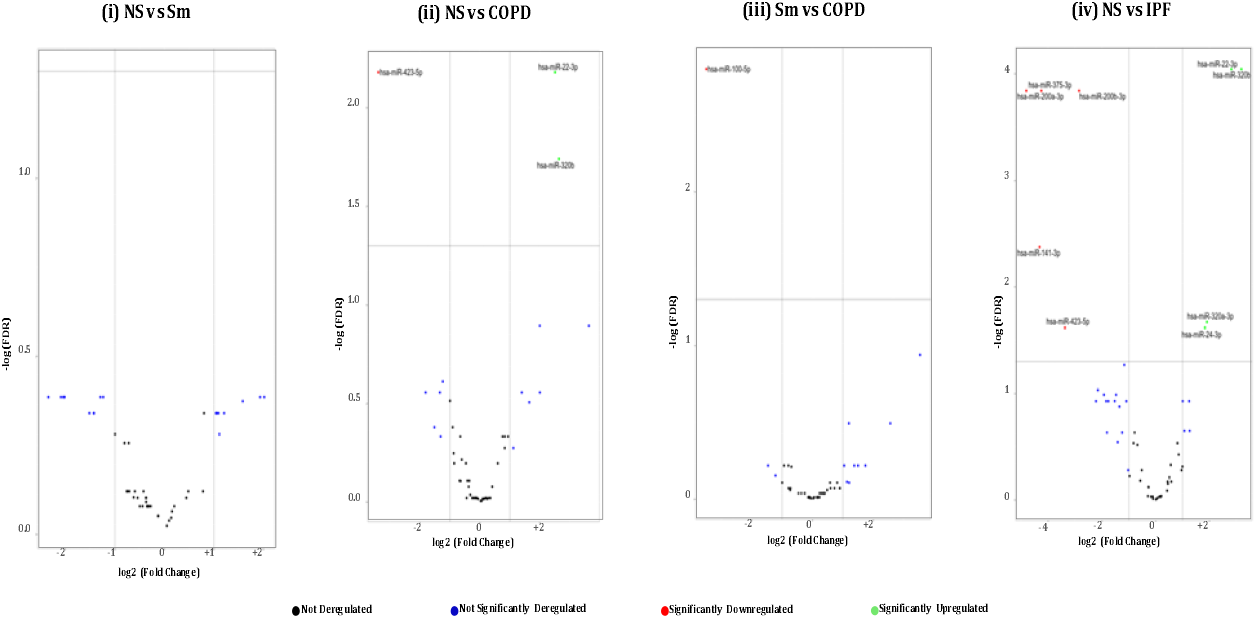
Volcano Plots showing number and distribution of miRNA from BALF-derived exosomes. Volcano plot showing the relation between –log(FDR) [Y-axis] vs log2(fold change) [X-axis] in the differentially expressed miRNAs amongst BALF exosomes derived from (i) healthy non-smokers (NS) vs healthy cigarette smokers (Sm), (ii) healthy non-smokers (NS) vs COPD patients (COPD), (iii) healthy cigarette smokers (Sm) vs COPD patients and (iv) healthy non-smokers (NS) and IPF patients (IPF).

**Figure 5:**
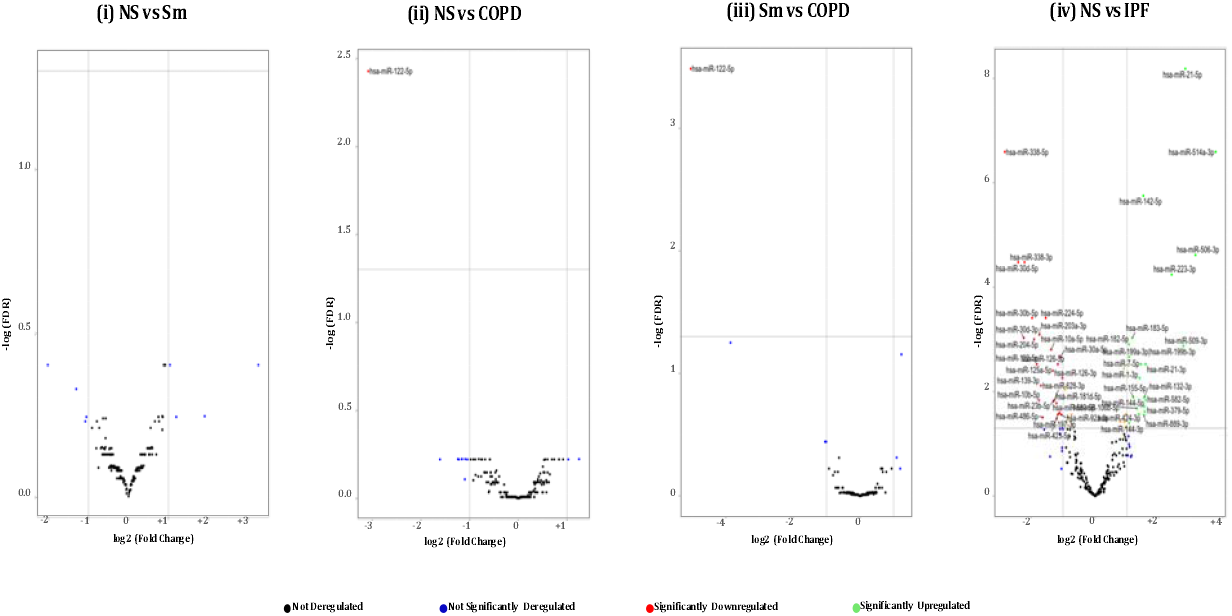
Volcano Plots showing number and distribution of miRNA from lung tissue-derived exosomes. Volcano plot showing the relation between –log(FDR) (Y-axis) vslog2(fold change) (X-axis) in the differentially expressed miRNAs amongst lung tissue exosomes derived from (i) healthy non-smokers (NS) vs healthy cigarette smokers (Sm), (ii) healthy non-smokers (NS) vs COPD patients (COPD), (iii) healthy cigarette smokers (Sm) vs COPD patients and (iv) healthy non-smokers (NS) and IPF patients (IPF).

### Hierarchical clustering identified differentially expressed miRNAs in the BALF or lung-derived exosomes from non-smokers, smokers, patients of COPD and IPF

We generated heat maps showing the top 50 differentially expressed miRNAs from the BALF and lung tissue-derived exosomes from NS vs Sm, NS vs COPD, NS vs IPF and Sm vs COPD as shown in Figs 6 &7. Each miRNA is depicted in the individual row of the heat map while the color scale represents the relative expression level as denoted in the scale bar alongside. A detailed information about the significantly altered miRNAs with their respective p-values and biological significance has been listed in Supplementary Tables 1&2. In brief, the following observations were made on comparing the various experimental pairs:

**Figure 6:**
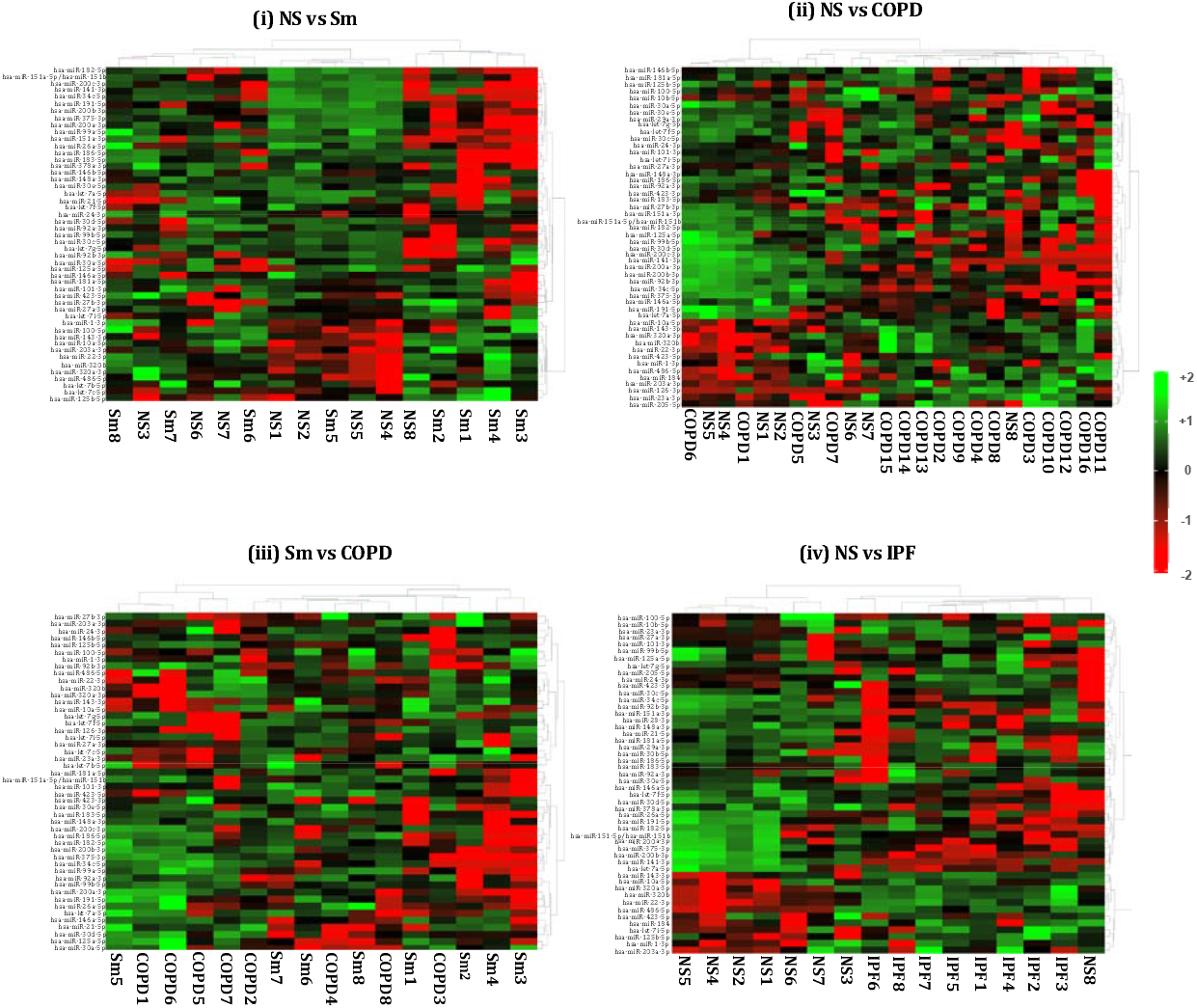
Hierarchical cluster analyses of differentially expressed miRNAs from BALF-derived exosomes. Heat map showing top 50 variable miRNAs that are differentially expressed in the BALF-derived exosomes from (i) healthy non-smokers (NS) vs healthy smokers (Sm), (ii) healthy non-smokers (NS) vs COPD patients (COPD), (iii) healthy smokers (Sm) and COPD patients (COPD) and (iv) healthy non-smokers (NS) and IPF patients (IPF).

**Figure 7:**
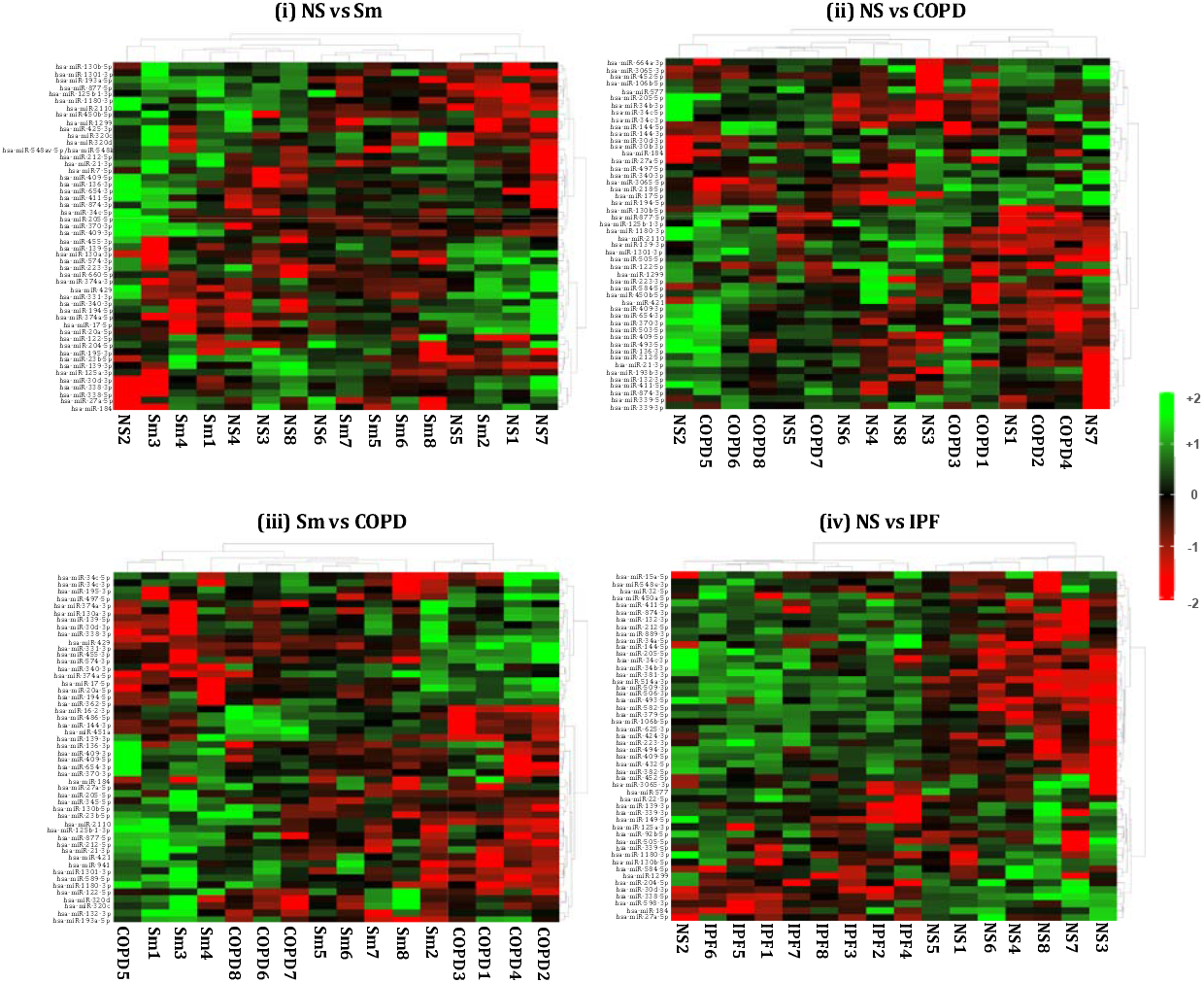
Hierarchical cluster analyses of differentially expressed miRNAs from lung tissue-derived exosomes. Heat map showing top 50 variable miRNAs that are differentially expressed in the lung tissue-derived exosomes from (i) healthy non-smokers (NS) vs healthy smokers (Sm), (ii) healthy non-smokers (NS) vs COPD patients (COPD), (iii) healthy smokers (Sm) and COPD patients (COPD) and (iv) healthy non-smokers (NS) and IPF patients (IPF)..

#### Non-smokers vs Smokers

We did not detect any significant differentially expressed miRNA in the BALF-derived exosomes from smokers and non-smokers. Similarly, no significant variation was observed on comparing the miRNA population from lung tissue-derived exosomes from smokers and non-smokers.

#### Non-smokers vs COPD

On comparing the BALF derived exosomal miRNAs from non-smokers and COPD patients, we found three significant differentially expressed miRNAs. Of these, two (miR-320b and miR-22-3p) were significantly upregulated, while one (miR-423-5p) was significantly downregulated in the BALF-derived exosomes from COPD patients as compared to the non-smoking controls. In contrast, we demonstrated significant downregulation of one (miR-122-5p) exosomal miRNA in the lung-tissues of COPD patients as compared to non-smokers.

#### Smoker vs COPD

We observed significant downregulation of miR-100-5p in the BALF-derived exosomes from COPD patients in comparison to those from healthy smokers.

Similarly, on comparing the lung-derived exosomes from these two study groups we found a significant downregulation of one miRNA. We noticed a significant downregulation of miR-122-5p in the exosomes derived from the lungs of COPD patients as compared to healthy smokers. Interestingly, the same miRNA was found to be downregulated on comparing the miRNA population from the lung-derived exosomes from COPD patients and non-smokers.

#### Non-smokers vs IPF

Our results showed a distinct miRNA signature in the BALF and lung tissue-derived exosomes from IPF patients as compared to non-smoking controls. A total of nine differentially expressed miRNAs were identified from the BALF-derived exosomes of IPF patients as compared to healthy non-smoking controls. Of the 9, five (miR-375-3p, miR-200a-3p, miR-200b-3p, miR-141-3p, and miR-423-5p) miRNAs were significantly downregulated; while four (miR-22-3p, miR-320a-3p, miR-320b, and miR-24-3p) were upregulated in the BALF of IPF patients.

Interestingly, we found 55 (26 upregulated and 29 downregulated) differentially expressed miRNAs in the lung-derived exosomes from lungs of IPF patients as compared to non-smoking controls.

### GO enrichment and KEGG analyses differentially expressed miRNAs from BALF and lung-derived exosomes in COPD and IPF patients

To understand the potential functions of the differentially expressed miRNAs in COPD and IPF patients, we performed GO enrichment covering three major domains-biological process, cellular compartment and molecular function. GO term annotation of differentially altered miRNAs in BALF-derived exosomes from COPD patients as compared to healthy non-smokers and smokers resulted in enrichment of terms including: post-translational protein modification, ubiquinone biosynthetic process, cellular component organization, membrane enclosed lumen, clathrin-coated vesicle, mitochondrial matrix, protein binding, protein heterodimerization and transferase activity. The regulatory pathway annotation by KEGG enrichment analyses showed representation of pathways involved in terpenoid backbone biosynthesis, cAMP signaling, cellular senescence and chemokine signaling amongst COPD patients. However, none of these pathways was significantly over-represented in our analyses. GO annotation for miRNA population obtained from IPF patient BALF resulted in enrichment of terms including, lipid transport, mesenchymal cell development, chromatin, mitochondria, lysosome, R-SMAD binding and ATPase activity. The KEGG analyses for this subject group showed 80% overlap with the pathways enriched amongst COPD patients. Interestingly, however, we found a significant overrepresentation of pathways regulating glycosaminoglycan biosynthesis (p=0.028) in the BALF-derived exosomes from IPF patients.

GO annotation of differentially regulated miRNAs from lung derived exosomes was conducted separately. We found enrichment of terms like, blood vessel morphogenesis, angiogenesis, transmembrane signaling receptor activity, G protein-coupled receptor activity, calcium mediated signaling and calcineurin-NFAT signaling cascade in lung-derived exosomes from COPD patients as compared to healthy individuals (non-smokers and smokers). Contrarily, GO terms including, plasma membrane bounded cell projection organization, chemical homeostasis, G protein-coupled receptor activity, positive regulation of phospholipase C activity, MHC class II protein complex signaling, GTPase activator activity, and positive regulation of non-membrane spanning protein tyrosine kinase activity were found to be enriched on analyzing differentially expressed miRNAs from lung-derived exosomes in IPF patients. KEGG enrichment analyses showed overrepresentation of pathways regulating apoptosis, asthma, and cGMP-PKG signaling pathway, amongst others in COPD patients. However, none of these regulatory pathways were significantly represented. Contrarily, KEGG enrichment analyses of miRNA profile from IPF patients identified representation of 40 pathways, of which 12 were significantly represented in the miRNA population from the lung-derived exosomes from IPF patients.

Tables 2-5 provide an account of the GO enrichment and KEGG analyses results for our comparisons of various subject groups in this study. Only selected pathways has been represented in the Tables.

**Table 2:**
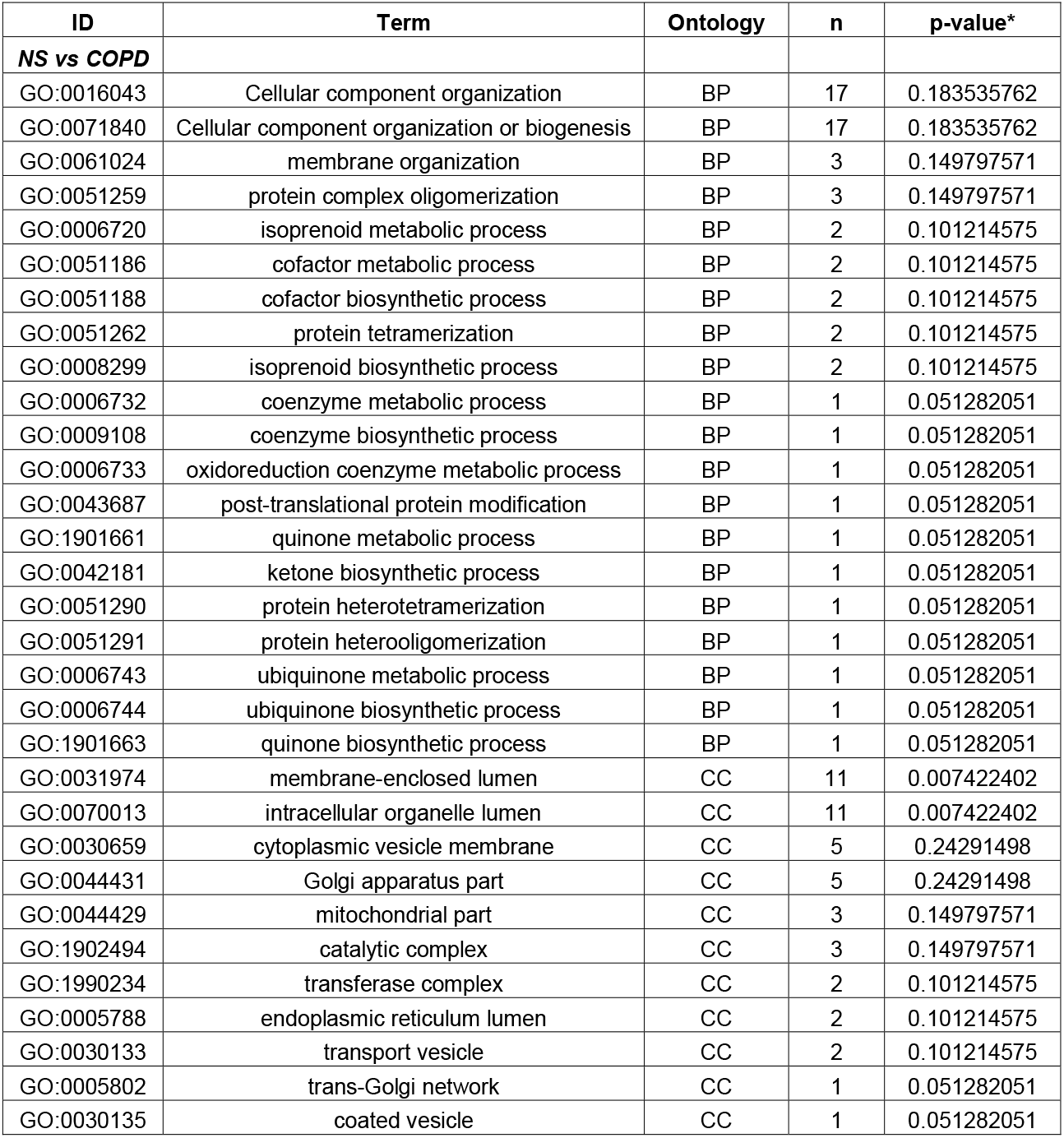
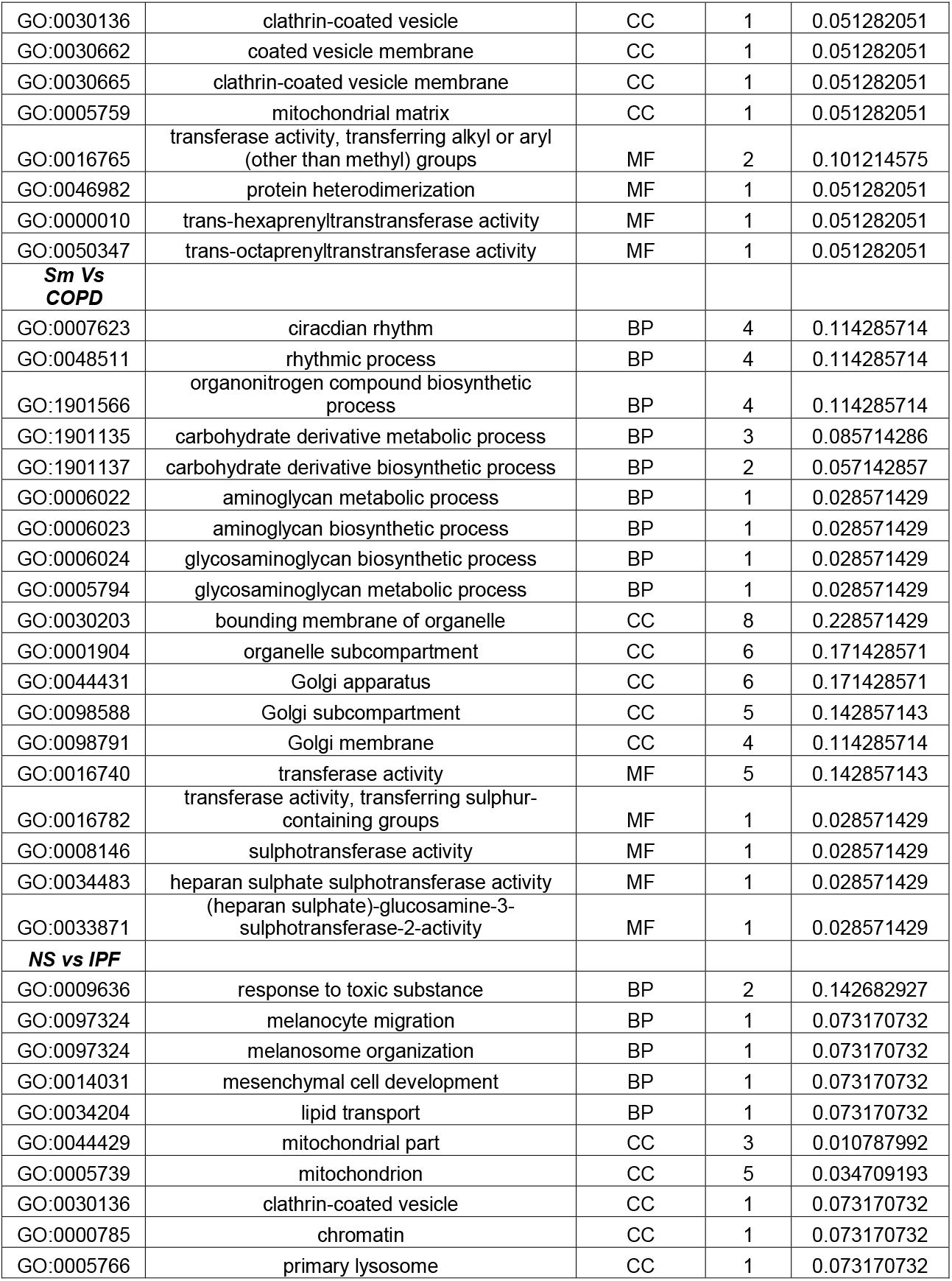
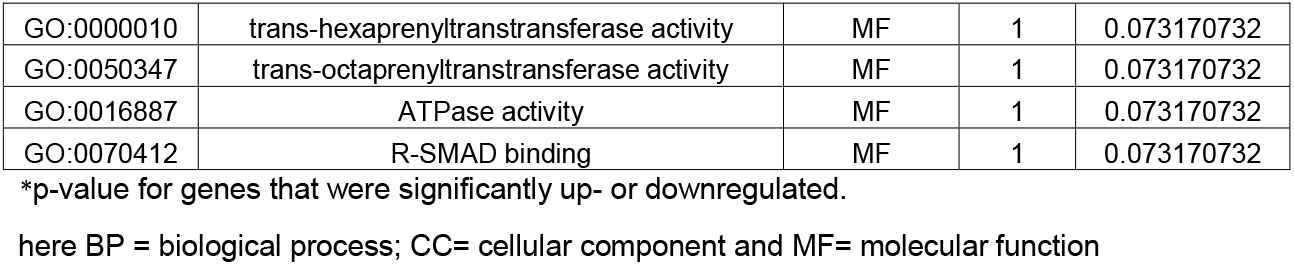
GO Enrichment Analysis of differentially expressed miRNAs in BALF-derived exosomes.

**Table 3:** GO Enrichment Analysis of differentially expressed miRNAs in Lung tissue-derived exosomes.

| ID | Term | Ontology | n | p-value* |
| --- | --- | --- | --- | --- |
| **NS vs COPD** |  |  |  |  |
| GO:0048514 | blood vessel morphogenesis | BP | 12 | 0.068181818 |
| GO:0050808 | synapse organization | BP | 11 | 0.0625 |
| GO:0051962 | positive regulation of nervous system development | BP | 11 | 0.0625 |
| GO:0044089 | positive regulation of cellular component biogenesis | BP | 10 | 0.056818182 |
| GO:0044430 | cytoskeletal part | CC | 9 | 0.051136364 |
| GO:0001525 | angiogenesis | BP | 8 | 0.045454545 |
| GO:0050803 | regulation of synapse structure or activity | BP | 8 | 0.045454545 |
| GO:0050807 | regulation of synapse organization | BP | 8 | 0.045454545 |
| GO:0015630 | microtubule cytoskeleton | CC | 8 | 0.045454545 |
| GO:0038023 | signaling receptor activity | MF | 8 | 0.045454545 |
| GO:0060089 | molecular transducer activity | MF | 8 | 0.045454545 |
| GO:0004888 | transmembrane signaling receptor activity | MF | 7 | 0.039772727 |
| GO:0004930 | G protein-coupled receptor activity | MF | 7 | 0.039772727 |
| GO:0019932 | second messenger-mediated signaling | BP | 6 | 0.034090909 |
| GO:0007416 | synapse assembly | BP | 6 | 0.034090909 |
| GO:0045765 | regulation of angiogenesis | BP | 5 | 0.028409091 |
| GO:1901342 | regulation of vasculature development | BP | 5 | 0.028409091 |
| GO:0051963 | regulation of synapse assembly | BP | 5 | 0.028409091 |
| GO:0051965 | positive regulation of synapse assembly | BP | 4 | 0.022727273 |
| GO:0019722 | calcium-mediated signaling | BP | 4 | 0.022727273 |
| GO:0005815 | microtubule organizing center | CC | 4 | 0.022727273 |
| GO:0005813 | centrosome | CC | 4 | 0.022727273 |
| GO:0016525 | negative regulation of angiogenesis | BP | 3 | 0.017045455 |
| GO:1901343 | negative regulation of vasculature development | BP | 3 | 0.017045455 |
| GO:2000181 | negative regulation of blood vessel morphogenesis | BP | 3 | 0.017045455 |
| GO:0033173 | calcineurin-NFAT signaling cascade | BP | 2 | 0.011363636 |
| GO:0048016 | inositol phosphate-mediated signaling | BP | 2 | 0.011363636 |
| GO:0097720 | calcineurin-mediated signaling | BP | 2 | 0.011363636 |
| **Sm Vs COPD** |  |  |  |  |
| GO:0048514 | blood vessel morphogenesis | BP | 11 | 0.071428571 |
| GO:0044087 | regulation of cellular component biogenesis | BP | 11 | 0.071428571 |
| GO:0044089 | positive regulation of cellular component biogenesis | BP | 10 | 0.064935065 |
| GO:0051962 | positive regulation of nervous system development | BP | 10 | 0.064935065 |
| GO:0001525 | angiogenesis | BP | 8 | 0.051948052 |
| GO:0050803 | regulation of synapse structure or activity | BP | 8 | 0.051948052 |
| GO:0050807 | regulation of synapse organization | BP | 8 | 0.051948052 |
| GO:0044430 | cytoskeletal part | CC | 7 | 0.045454545 |
| GO:0038023 | signaling receptor activity | MF | 7 | 0.045454545 |
| GO:0060089 | molecular transducer activity | MF | 7 | 0.045454545 |
| GO:0019932 | second-messenger-mediated signaling | BP | 6 | 0.038961039 |
| GO:0007416 | synapse assembly | BP | 6 | 0.038961039 |
| GO:0015630 | microtubule cytoskeleton | CC | 6 | 0.038961039 |
| GO:0004888 | transmembrane signaling receptor activity | MF | 6 | 0.038961039 |
| GO:0045765 | regulation of angiogenesis | BP | 5 | 0.032467532 |
| GO:1901342 | regulation of vasculature development | BP | 5 | 0.032467532 |
| GO:0051963 | regulation of synapse assembly | BP | 5 | 0.032467532 |
| GO:0051965 | positive regulation of synapse assembly | BP | 4 | 0.025974026 |
| GO:0019722 | calcium-mediated signaling | BP | 4 | 0.025974026 |
| GO:0016525 | negative regulation of angiogenesis | BP | 3 | 0.019480519 |
| GO:1901343 | negative regulation of vasculature development | BP | 3 | 0.019480519 |
| GO:2000181 | negative regulation of blood vessel morphogenesis | BP | 3 | 0.019480519 |
| GO:0005815 | microtubule organizing center | CC | 3 | 0.019480519 |
| GO:0005813 | centrosome | CC | 3 | 0.019480519 |
| GO:0004930 | G-protein coupled receptor activity | MF | 3 | 0.019480519 |
| GO:0033173 | calcineurin-NFAT signaling cascade | BP | 2 | 0.012987013 |
| GO:0048016 | inositol phosphate-mediated signaling | BP | 2 | 0.012987013 |
| GO:0097720 | calcineurin-mediated signaling | BP | 2 | 0.012987013 |
| **NS Vs IPF** |  |  |  |  |
| GO:0065008 | regulation of biological quality | BP | 44 | 0.0327192 |
| GO:0007399 | nervous system development | BP | 36 | 0.006858696 |
| GO:0048878 | chemical homeostasis | BP | 18 | 0.003415 |
| GO:0030030 | cell projection organization | BP | 17 | 0.012599483 |
| GO:0120036 | plasma membrane bounded cell projection organization | BP | 17 | 0.01259943 |
| GO:0044459 | plasma membrane region | CC | 10 | 0.028105097 |
| GO:0007416 | synapse assembly | BP | 7 | 0.006212841 |
| GO:0030424 | axon | CC | 7 | 0.0488352 |
| GO:0150034 | distal axon | CC | 6 | 0.0305685 |
| GO:0031349 | positive regulation of defense response | BP | 5 | 0.016747 |
| GO:0044306 | neuron projection terminus | CC | 4 | 0.007341699 |
| GO:0008092 | cytoskeletal protein binding | MF | 4 | 0.0073417 |
| GO:0004930 | G-protein coupled receptor activity | MF | 4 | 0.083811139 |
| GO:0051965 | positive regulation of synapse assembly | BP | 4 | 0.007341699 |
| GO:0010863 | positive regulation of phospholipase C activity | BP | 2 | 0.01653348 |
| GO:0043235 | receptor complex | CC | 2 | 0.01653348 |
| GO:0023026 | MHC class II protein complex binding | MF | 2 | 0.0165335 |
| GO:0005096 | GTPase activator activity | MF | 2 | 0.0165335 |
| GO:1903997 | positive regulation of non-membrane spanning protein tyrosine kinase activity | BP | 2 | 0.01653348 |

\*p-value for genes that were significantly up- or downregulated.
here BP = biological process; CC= cellular component and MF= molecular function

**Table 4:**
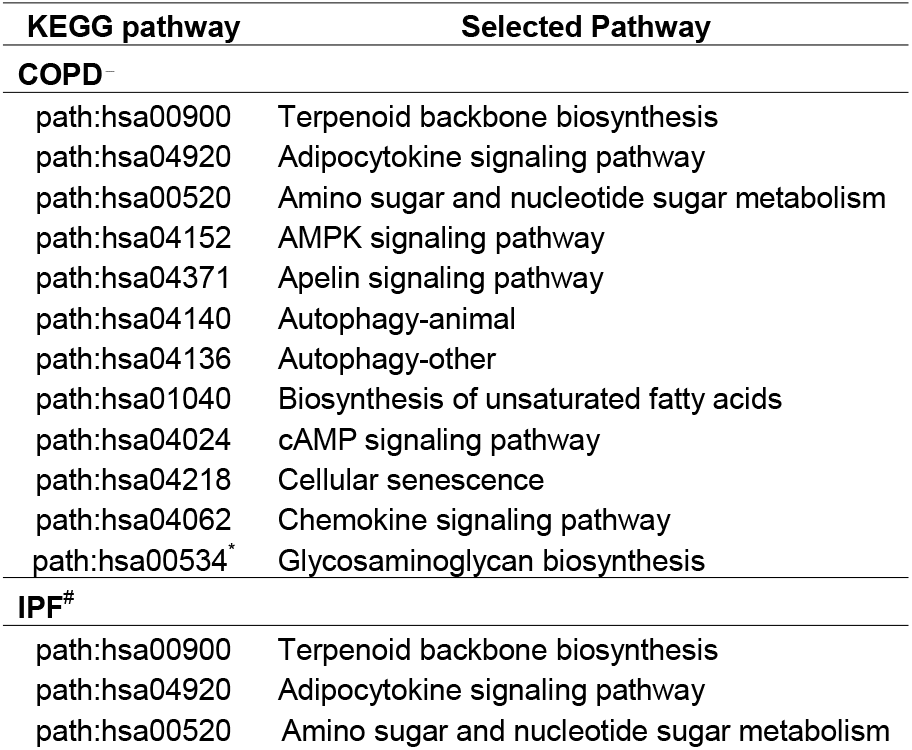
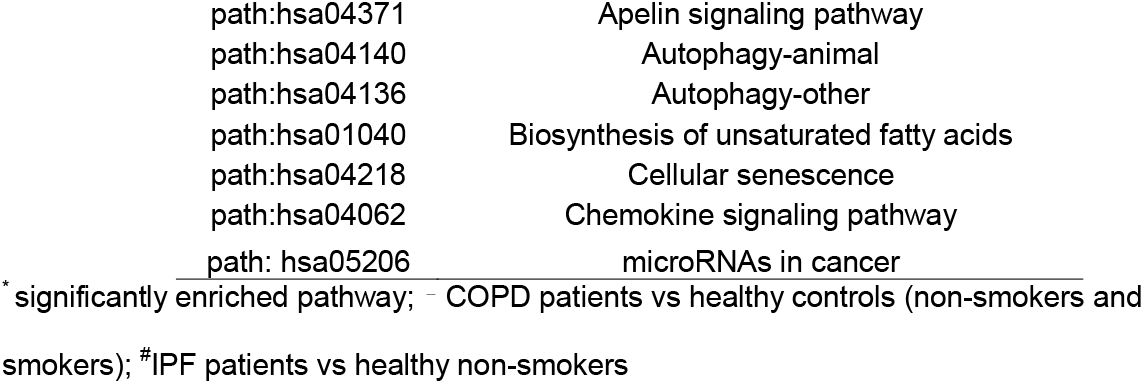
KEGG Analyses of differentially expressed miRNAs in BALF-derived exosomes from COPD and IPF patients.

**Table 5:**
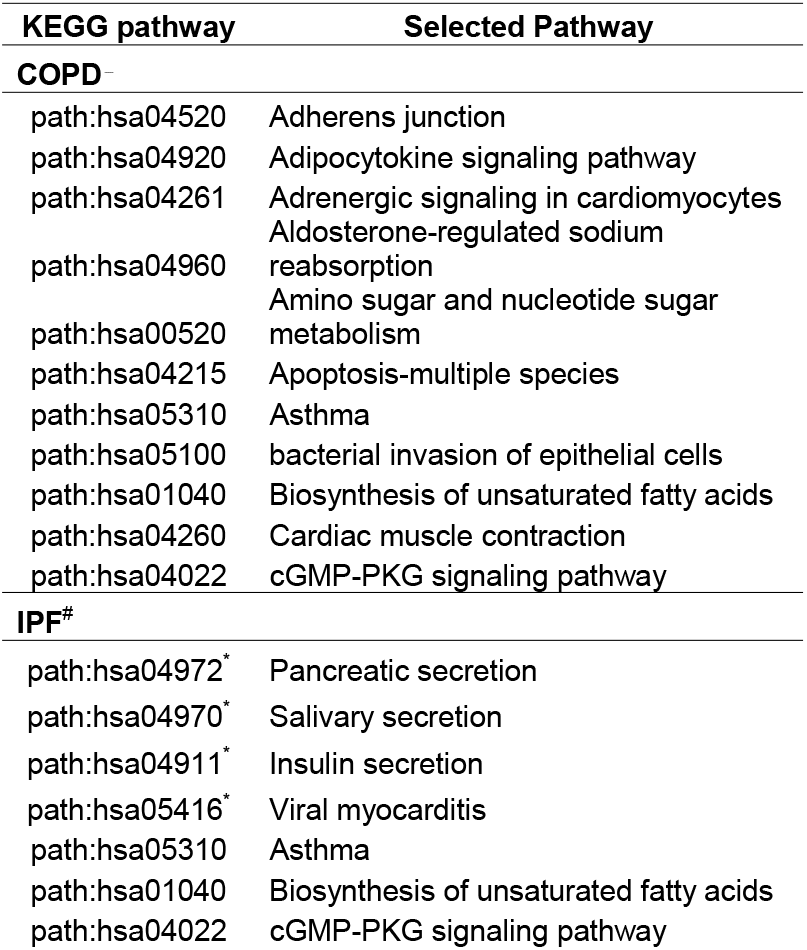
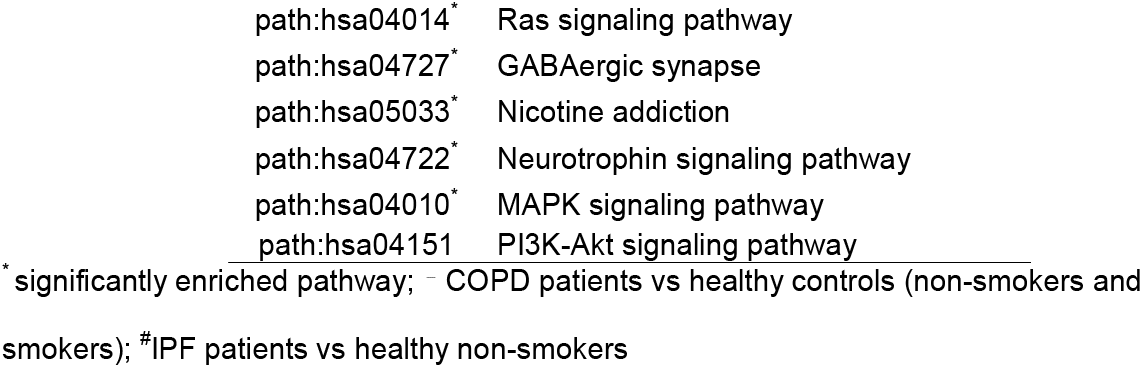
KEGG Analyses of differentially expressed miRNAs in lung tissue-derived exosomes from COPD and IPF patients.

## Discussion

The role of exosomes in lung diseases has gained increasing attention in recent times due to their role in influencing intercellular communication. These are 50-150 nm in diameter, membrane-bound vesicles that contain protein, DNA, mRNA, microRNA (miRNA) and small non-coding RNAs to regulate pleiotropic functions(41). Recent studies suggest that exosomes mediate cellular crosstalk in lung microenvironment and that cigarette smoke-induced exosomes promote myofibroblast differentiation in primary lung fibroblasts (21, 22). In addition, activated exosomes (due to cigarette smoke or disease conditions) result in macrophage polarization and matrix destruction in mouse models (42, 43). These studies implicate that exosomes affect cell-to-cell signaling in tobacco smoke-related disorders.

In this respect, inhalation of toxic agents from tobacco smoke might result in irreparable airway injury leading to various lung diseases like COPD and IPF. While the etiology/cause of each of these diseases might be environmental factors, the disease pathologies are distinct (41). Therefore, we were interested in understanding if the exosomal population and the exosome-derived miRNA signatures from BALF and lung tissues of non-smokers, smokers, COPD patients and IPF patients are unique and can be developed into effective biomarkers for the clinical diagnosis of respective pathologies.

Results from next generation sequencing revealed no significant differentially expressed miRNAs in the BALF or lung-derived exosomes from heathy smokers and non-smokers. This suggests that smoking status alone does not affect the exosome-mediated signaling in healthy individuals. However, we found a distinct variation in the miRNA populations from BALF and lung tissue-derived exosomes from COPD patients in comparison to healthy non-smokers. We found a 3-fold downregulation in the expression of miR-423-5p in the BALF-derived exosomes from COPD patients as compared to healthy non-smoking controls. Of note, miR-423-5p is known to be involved in the regulation of apoptosis and extracellular matrix degradation in human nucleus pulposus cells (44). Contrary to our findings, Molina-Pinelo *et al* (2014) identified increased expression of miR-423-5p in the BALF collected from COPD patients as compared to the control group. However, it is important to mention here that the control group included in this study comprised of a few ex-smokers and they did not look at the exosome-derived miRNA from BALF (45). So taken together, it can be concluded that miR-423-5p is crucial in COPD and must be studied further to understand its potential role in the pathophysiology of COPD. Further, we observed two-fold increase in the expression of miR-320b and miR-22-3p in the BALF-derived exosomes from COPD patients as compared to the non-smoking controls. Previous study by our group identified upregulation of both miR-320b and miR-22-3p in the peripheral blood-derived exosomes of COPD patients (11), thus indicating significant role of these miRNAs in regulating the disease phenotype. miR-320b is the negative regulator of mitochondrial mediator, TP53-regulated inhibitor of apoptosis (TRIAP1), and has been previously shown to be upregulated in the peripheral blood nuclear cells (PBMCs) from COPD patients (46, 47). Similarly, miR-22-3p is reported to inhibit HDAC4 to promote Th17-mediated emphysema in cigarette smoke (4 month)-exposed C57Bl/6 mice lungs (48). Serum levels of miR-22-3p are known to be increased amongst COPD patients based on their history of smoking, thus revealing the crucial nature of this miRNA in the progression of the disease (49).

On comparing the miRNA expression of lung tissue-derived exosomes from COPD patients and non-smokers, we observed 3-fold downregulation of miR-122-5p in the lungs of COPD patients as compared to healthy non-smoking controls. Importantly, we further observed a 5-fold decrease in the expression of miR-122-5p on comparing miRNA population from lung-derived exosomes from COPD patients versus healthy smokers. Our results are in accordance with previous literature (50-52). For instance, Zhu *et al* (2020) demonstrated the downregulation of miR-122-5p in the sputum and plasma of COPD patients and proved that it functions as a negative regulator of IL-17A production (50). It is pertinent to mention here that though we did not find any commonly altered miRNAs in the exosomes from BALF or lung tissues of COPD patients, we found links that associate miRNA-mediated modulation of IL17-signalling amongst the diseased individual. The role of IL-17 in the disease pathology of COPD is rapidly emerging and is known to play an important role in the regulation of chronic inflammation and emphysema in COPD (53). Hence, our findings identify the upstream regulators of this pathway that could possibly alter the IL17-mediated inflammation in patients with advancing disease.

Next, we identified significant downregulation of miR-100-5p in the BALF-derived exosomes from COPD patients as compared to healthy smokers. Functionally, miR-100 has been linked to the regulation of epithelial-mesenchymal transition (EMT), apoptosis and inflammation (54, 55). Furthermore, Akbas and colleagues have demonstrated downregulation of miR-100-5p in the serum of COPD patients when compared to healthy smokers, which is in accordance to our study results (56).

The differentially expressed miRNA population from BALF and lung tissue-derived exosomes in COPD and IPF was very distinct in our study. We found five significantly downregulated (miR-200a-3p, miR-200b-3p, miR-141-3p, miR-375-3p, and miR-423-3p) whereas four significantly upregulated (miR-320a-3p, miR-320b, miR-22-3p and miR-24-3p) miRNAs in the BALF-derived exosomes from IPF patients. Of these, miR-423-3p and miR-320b were found to be significantly dysregulated amongst COPD patients as well. Of note, existing reports suggest a role of miR-200 in the pathogenesis of IPF (57, 58). It has been shown that miR-200 promotes TGF-β1-induced EMT in normal cells and its downregulation results in fibrogenic phenotype in IPF (57). To our knowledge, there is no existing literature associating miR-141-3p, miR-22-3p and miR-24-3p with IPF. Thus, we for the first time identify the association of these miRs with the disease pathogenesis in IPF.

We found 55 differentially expressed miRNAs in the lung-derived exosomes from IPF patients when compared to non-smokers. Of these, many including miR514-3p, miR-122-5p, miR-10b-5p, miR-139b-3p, miR-582-5p, miR-889-3p, miR-1-3p, miR-148a-3p and miR-151b, have never been reported with IPF. Our study for the first time reports the correlation of the dysregulated expression of these miRNAs in the lung derived exosomes from IPF patients. Of note, we observed a three-fold increase in the expression of miR-506-3p in the lung-derived exosomes from IPF patients as compared to the healthy non-smoking controls. Previous work by Zhu et al (2019) reported that miR-506-3p targets p65 subunit of NF-κB to induce apoptosis and inflammation in experimental mice model for IPF. This study concluded that miR-506-3p is a regulator of lung fibrosis (59). Our results provide clinical evidence suggesting a crucial role of this miRNA in the pathophysiology of IPF in humans. Similarly, accumulating evidence support the role of miR-21-5p in the disease progression of IPF (60-62). Further, the expression of miR-21-5p is controlled by the levels of TGF-β family proteins and SMADs, both of which are key regulators in the etiology of fibrosis (63).

Our study had some limitations. Firstly, the sample size for each of the study groups was quite small (n=8-16). In addition, due to non-availability of age- and gender-matched individuals in our cohorts, we were unable to normalize for the gender and age-specific bias in our results. Further limitations were the non-availability of non-smokers/never-smokers and limited information regarding the spirometry, pack-years and smoking history of all the subjects included in this study, which may have affected the final interpretation of our findings.

## Conclusions

Overall, this is the first study that compares the BALF and lung tissue-derived exosomal miRNAs from IPF and COPD patients with healthy subjects to suggest the unique miRNA signatures that could develop as a biomarker to identify the disease progression of these pulmonary conditions. Future studies will be designed to validate the findings from this work and to understand the role of exosomal miRNAs in affecting the disease development, progression and severity in COPD and IPF.

## Data Availability

All data are available online.
: All the data included in this manuscript is available online and is free to access to all readers. The NGS data and/or analyzed files during the current study are available at Gene Expression Omnibus accession number GSE180651 (https://www.ncbi.nlm.nih.gov/geo/query/acc.cgi?acc=GSE180651). Data available date: August 15-2021.

## List of Abbreviations

COPD: Chronic Obstructive Pulmonary disease
IPF: Idiopathic Pulmonary disease
TEM: Transmission Electron Microscopy
NGS: Next Generation sequencing miRNA: micro RNA
BALF: Bronchoalveolar Lavage fluid
EMT: epithelial-mesenchymal transition

## Acknowledgements

The authors would like to acknowledge Drs. Isaac Sundar and Dongmei Li for their scientific inputs. We would further like to thank Sarah McNamara, research nurse at Centre for Inflammation Research, University of Edinburgh for providing patient care during sample collection for this study. We would like to acknowledge Dr. Robert Foronjy, Associate Professor at SUNY Downstate Health Sciences University for for clinical support for some of the BALF samples included in the study. We would also like to acknowledge Dr. Ashokkumar Srinivasan for his assistance in optimization of study protocol.

## Declarations

The authors have declared that no competing interests exist.

## Funding

The National Institutes of Health (NIH) HL137738, HL133404 and HL135613 supported this work.

### Disclaimer

None

### Meeting Presentation

None

### Additional Contributions

None

### Data Availability

All the data included in this manuscript is available online and is free to access to all readers. The NGS data and/or analyzed files during the current study are available at Gene Expression Omnibus accession number GSE180651 (https://www.ncbi.nlm.nih.gov/geo/query/acc.cgi?acc=GSE180651). Data available date: August 15-2021.

### Availability of data and materials

All authors confirm the availability of data and materials online/free access to readers.

### Authors Contribution

GK and KPM designed and conducted the experiments, GK and IR wrote, edited and/or revised the manuscript. GK was responsible for data curation, IR conceptually designed the overall experiments and manuscript, and acquired funding. MC, HSC, FL, NK, MAH provided BALF and/or lung tissues and edited the manuscript.

---

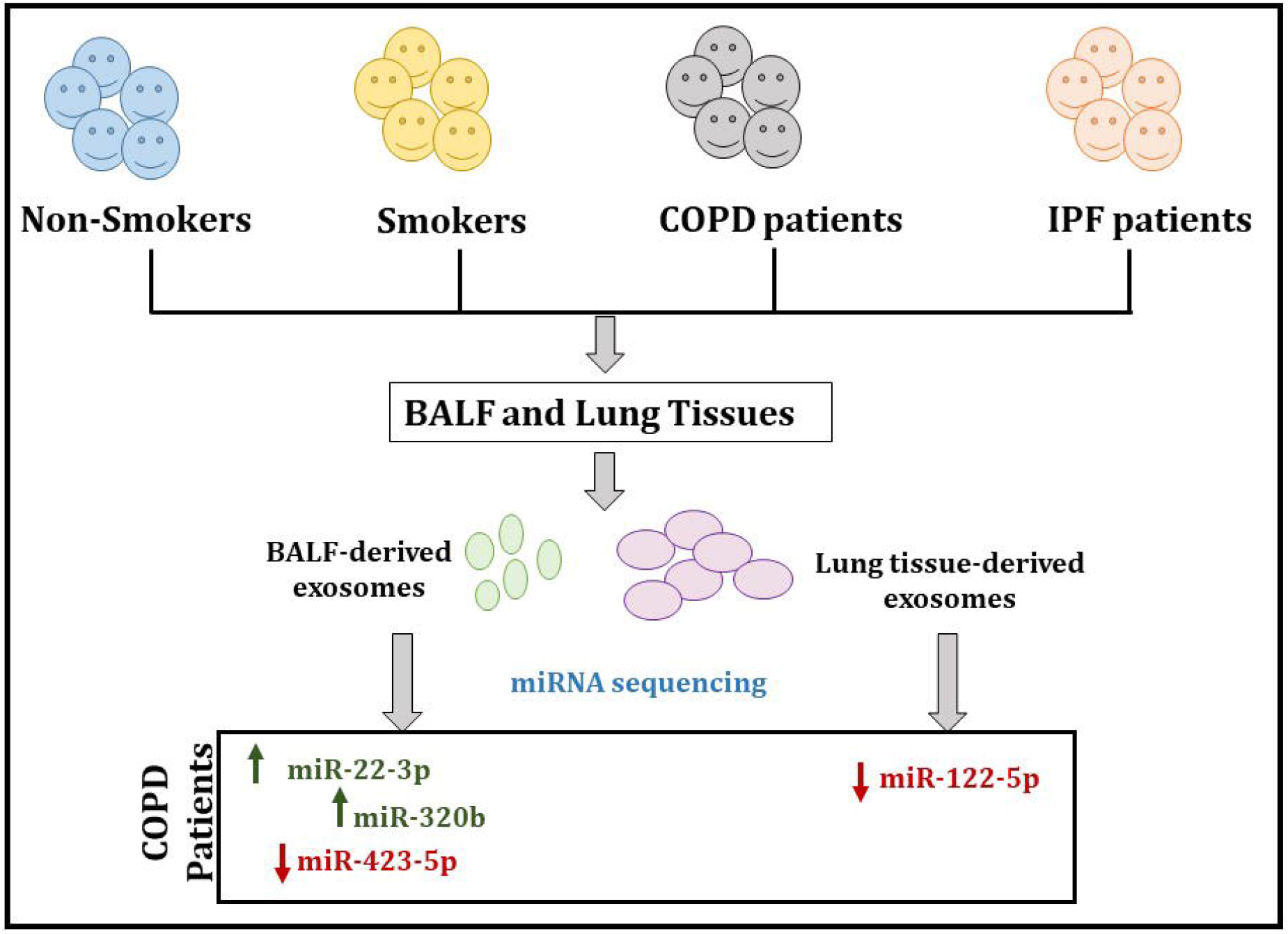

