## Supplementary material for "Distinct exosomal miRNA profiles from BALF and lung tissue from COPD and IPF patients": Suppl Figures

### Uncut Original Blots

<u>Full unedited gel for Figure 1(iii)</u>

Exosome Markers CD9 and CD81 in BALF exosomes

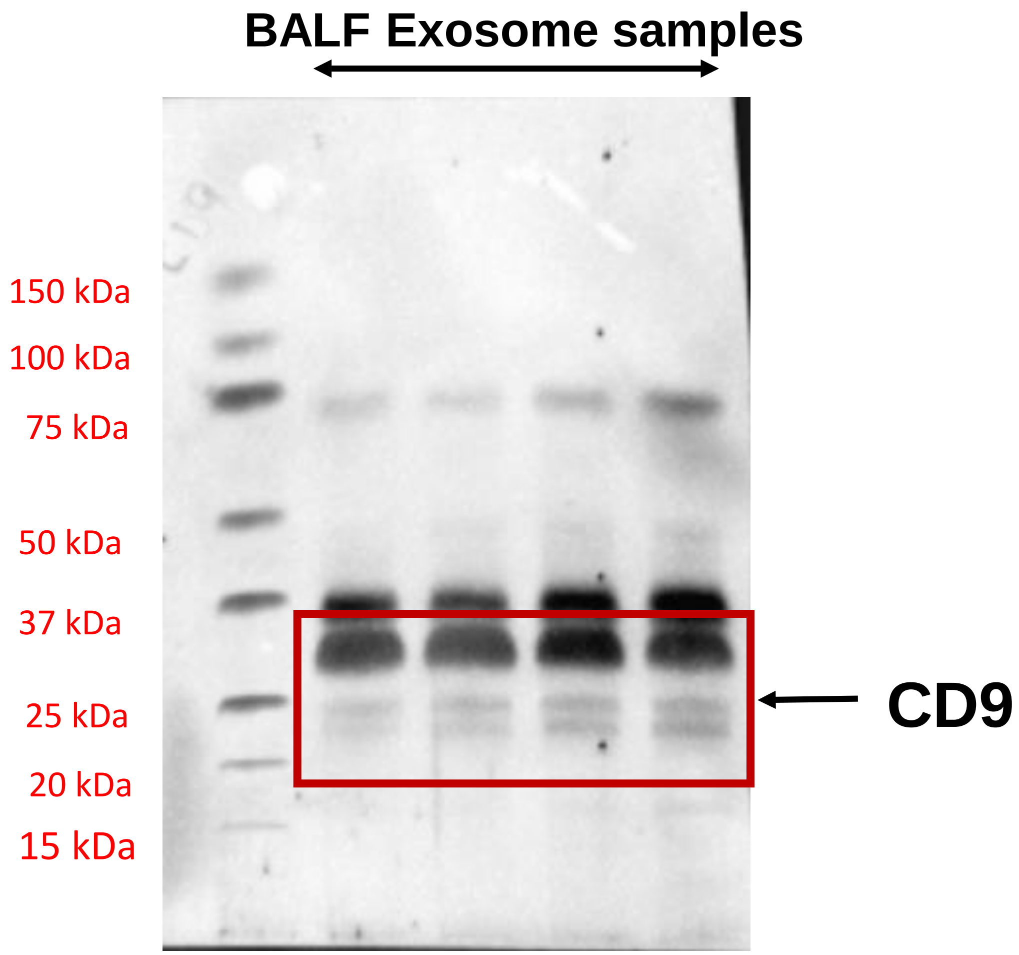


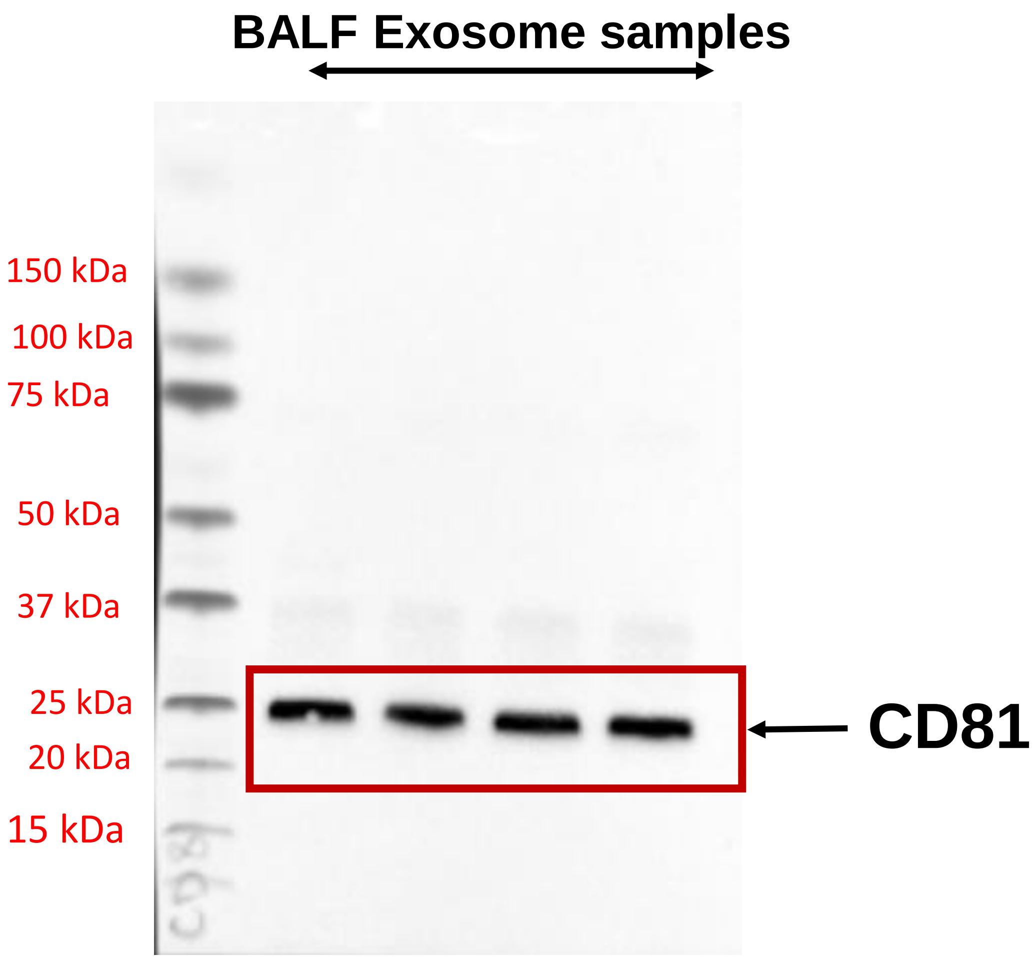


This shows the original uncut blots for CD9 and CD81 as shown in Figure 1(iii) of the manuscript

### Uncut Original Blots

<u>Full unedited gel for Figure 2(iii)</u>

Exosome Markers CD63 and CD81 in Lung tissue exosomes

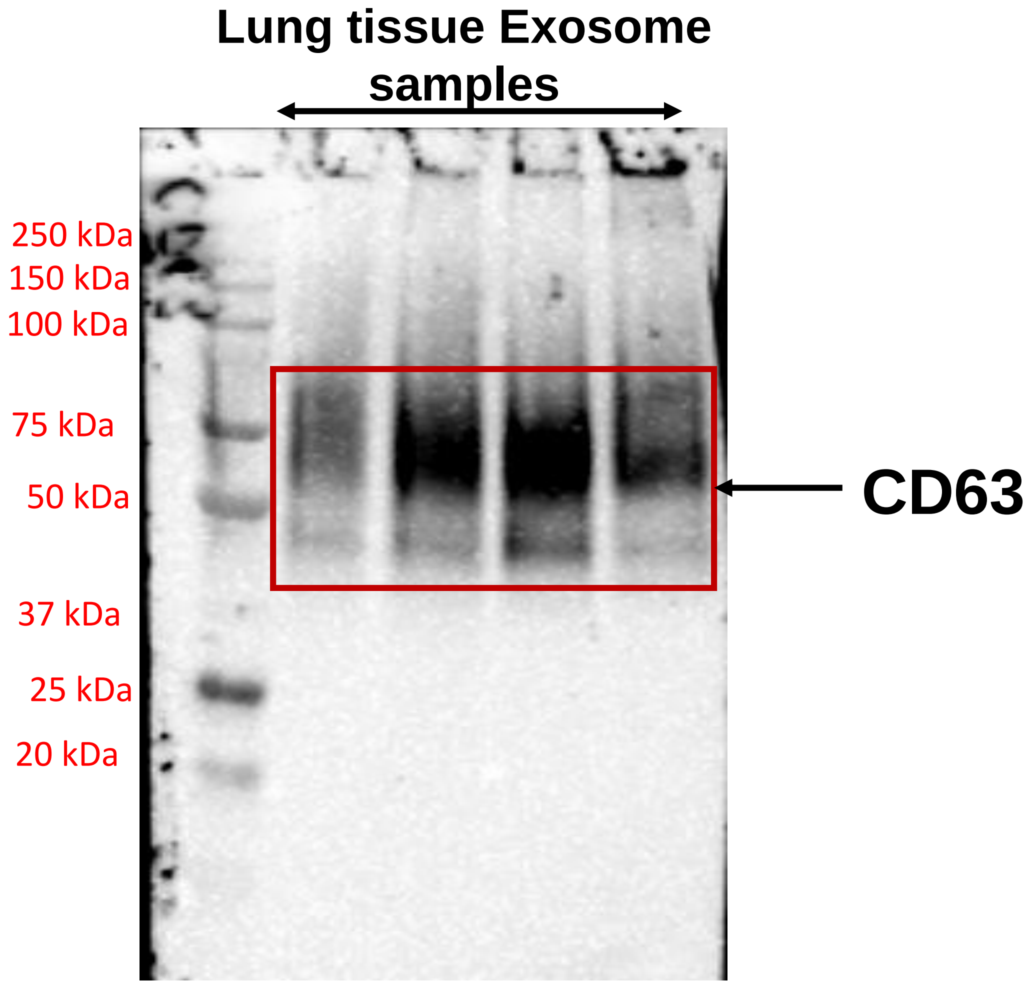


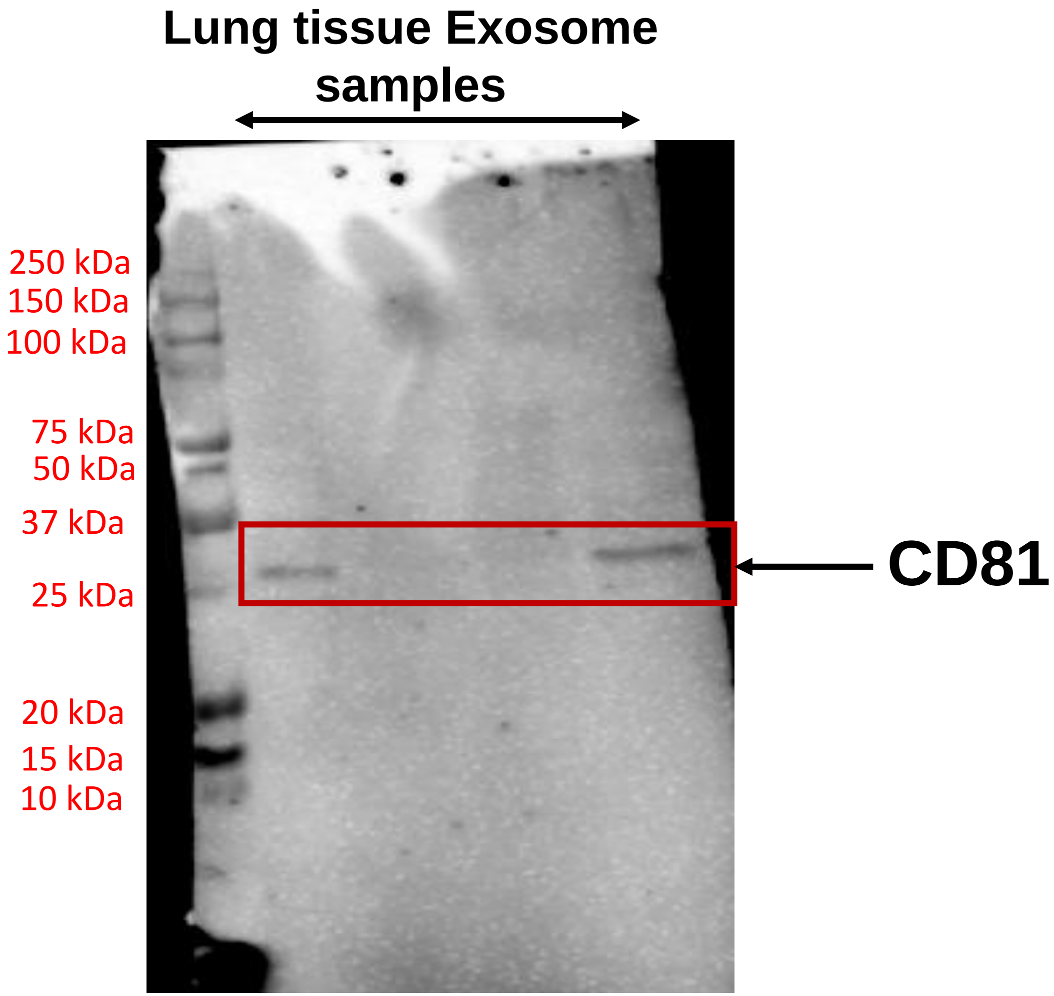


This shows the original uncut blots for CD9 and CD81 as shown in Figure 2(iii) of the manuscript
